# Comparative evaluation of the transmissibility of SARS-CoV-2 variants of concern

**DOI:** 10.1101/2021.06.25.21259565

**Authors:** Liang Wang, Xavier Didelot, Yuhai Bi, George F Gao

## Abstract

Since the start of the SARS-CoV-2 pandemic in late 2019, several variants of concern (VOC) have been reported, such as B.1.1.7, B.1.351, P.1, and B.1.617.2. The exact reproduction number *R*_t_ for these VOCs is important to determine appropriate control measures. Here, we estimated the transmissibility for VOCs and lineages of SAR-CoV-2 based on genomic data and Bayesian inference under an epidemiological model to infer the reproduction number (*R*_t_). We analyzed data for multiple VOCs from the same time period and countries, in order to compare their transmissibility while controlling for geographical and temporal factors. The lineage B had a significantly higher transmissibility than lineage A, and contributed to the global pandemic to a large extent. In addition, all VOCs had increased transmissibility when compared with other lineages in each country, indicating they are harder to control and present a high risk to public health. All countries should formulate specific prevention and control policies for these VOCs when they are detected to curve their potential for large-scale spread.

## Introduction

As the seventh coronavirus which could infect humans and then caused Coronavirus Disease 2019 (COVID-19), SARS-CoV-2 (also known as 2019-nCoV, or HCoV-19)^1^ was first identified in Wuhan China, in late 2019^2-4^. Within a few weeks, SARS-CoV-2 spread all over the world, and caused a global pandemic^5^ declared by the World Health Organization (WHO), which is the only pandemic caused by a coronavirus to date. As of 8^th^ June 2020, there are more than 172 million confirmed cases from more than 200 countries with more than three million deaths^6^, posing a global threat to public health. Furthermore, the global spread of COVID-19 has also thoroughly taxed the medical systems and global economies.

The transmissibility of infectious diseases can be measured by the basic reproduction number *R*_0_, which indicates how many secondary infectees could, on average, be directly caused by one infector in a susceptible population. The higher the *R*_0_, the higher the transmissibility of the infectious disease, which also means that the infectious disease is more difficult to be controlled. By extension, the temporal reproduction number *R*_t_ can be defined as the average number of secondary infections at time t. Traditionally, *R*_t_ is estimated using epidemiological data, which can be either individual contact tracing data or population-scale incidence data fitted with systems of ordinary differential equations and that represents a population-level epidemiological model^7^. However, getting unbiased datasets to apply these methods can be challenging. Here we used an alternative which is to use sequencing data to reconstruct a transmission tree which is informative about *R*_t_. As previous described, mutations in the genome of the SARS-CoV-2 have frequently occurred and accumulated during the epidemic. Some of these mutations may have increased the transmissibility, whereas the majority would likely have had no effect, but are still useful to reconstruct transmission trees. The assessment of the effect of mutations on transmissibility has been mainly based on non-human experimental animals (like hamsters *etc*), and it is still controversial whether these conclusions apply to humans. Besides, the timely adjustment of epidemic prevention and control strategies also requires a rapid assessment of the impact of newly emerging important mutations within pathogens’ genomes on transmission. Furthermore, several types of SARS-CoV-2 variants of concern (VOC) emerged during the pandemic, such as B.1.1.7 (WHO label: Alpha), B.1.351 (WHO label: Beta), P.1 (WHO label: Gamma), and B.1.617.2 (WHO label: Delta) *etc*. Under these circumstance, novel methods are needed that can quickly evaluate the impact of mutations on transmissibility.

Here, we estimated *R*_t_ for different lineages of SARS-CoV-2 based on genomic data and Bayesian inference under an epidemiological model, and then inferred the offspring distribution. The mean of the offspring distribution is the temporal reproductive number *R*_t_, which depends on both the pathogen transmissibility and the conditions in the host population (for example the proportion of immunized individuals or the control measures in place). To account for this, we compared the *R*_t_ of different lineages in the same country and during same periods to quantify the difference transmissibility between different lineages, especially for the previous described VOCs.

## Results

### Lineage B has a higher transmissibility than lineage A

Since only the United States and Australia contained sufficient numbers of viral genomes from both lineage A and B during the early phase of the COVID-19 pandemic, we used data from these two countries to compare the transmissibility between lineages A and B. The mean *R*_t_ for lineage A from Australia and USA were estimated as 1.75 (95% credible intervals (CI) 1.43-2.11) and 1.74 (95% CI 1.61-1.89), respectively (Figure 1A). However, the mean *R*_t_ for lineage B from Australia and USA were estimated as 2.33 (95% CI 2.05-2.64) and 3.18 (95% CI 2.76-3.63), respectively (Figure 1A). Firstly, the *R*_t_ of lineage B is significantly greater than that of lineage A, indicating higher transmissibility of lineage B compared to lineage A. This might be the reason why strains from lineage B rapidly became dominantly all over the world (Figure 1B). Secondly, the *R*_t_ of lineage A from the two countries are very close, however, the *R*_t_ of lineage B varied greatly between Australia and USA. We then found that the composition of lineage was significantly different between the datasets from these two countries (Figure 1C and D, *p*<0.01, Fisher’s exact test, two-sided). We speculated that different sub-lineages within lineage B might have different transmissibility and then tested the hypothesis by conducting further analysis. Since the data from lineage A was limited, the evaluation of transmissibility for each sub-lineage was mainly focused on those from lineage B and other emerging lineages in the same country during the same periods. In order to reduce the amount of calculation but at the same time be able to test the above hypothesis, only the dominant lineages showing exponential growth in each country were selected to perform the further analysis and comparison.

**Figure 1.**
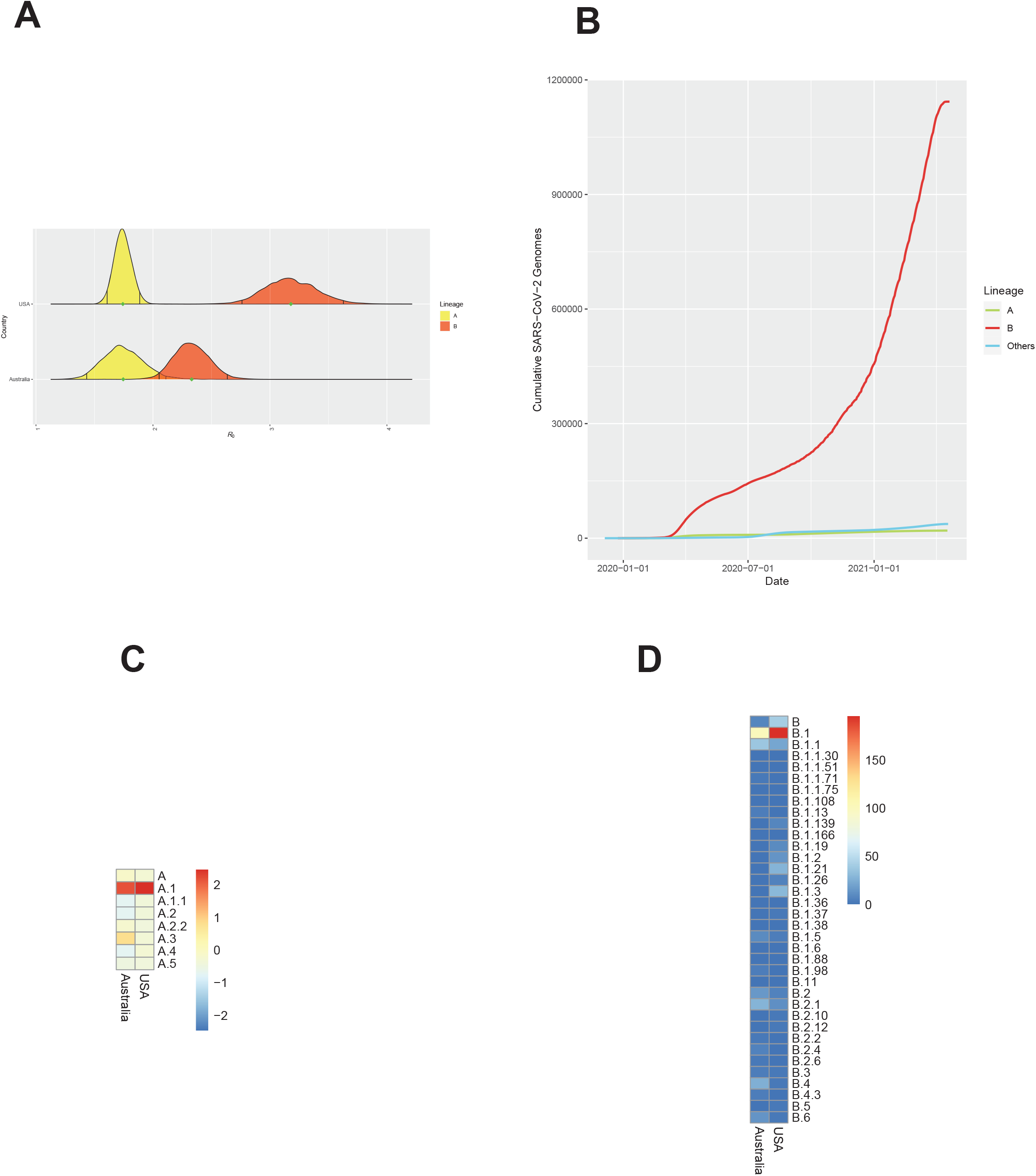
Difference in transmissibility between lineages A and B. A. The distribution of *R*_t_ for each lineage. The black line in each distribution indicated the 95% CI. B. The cumulative number of SARS-CoV-2 genomes for each lineage all over the world. C. The heatmap of number of viral genomes for each sub-lineage in lineage A. D. The heatmap of number of viral genomes for each sub-lineage in lineage B.

### B.1.1.7 has a higher transmissibility than other dominant lineages in UK

The composition of lineages in UK is shown in Figure 2A. B.1.177 was the dominant strain before 2021. We also found that the number of viral genomes from England far exceeds that from other parts of the UK (Figure 2B). Besides, according to the accumulation of number of viral genomes from each lineage in England, we could find that only three lineages (B.1.177, B.1.1.37, B.1.1.7) grew exponentially after October 2020 (Figure 2B). Taken together, only transmissibility of these three lineages were evaluated during October 2020 to January 2021 in this study, so that the impact of non-pharmaceutical interventions on the estimation of *R*_t_ will be consistent for different lineages. The *R*_t_ for B.1.177, B.1.1.37, B.1.1.7 were estimated as 1.08 (95% CI 1.072-1.09), 1.068 (95% CI 1.05-1.086), and 1.186 (95% CI 1.158-1.213) (Figure 2C). The B.1.177, B.1.1.37 had similar *R*_t_ which were both close to 1. However, B.1.1.7 had a significantly higher transmissibility than these two lineages. We next tested if the significantly high *R*_t_ could be affected by sampling bias. After five independently repeated sampling and subsequent analysis, we found that all these *R*_t_ for B.1.1.7 were close to each other, ranging from 1.178 to 1.194. Besides, all the 95% credible intervals from repeated sampling also did not have any intersection with those from lineage B.1.177 and B.1.1.37. Thus, the sampling bias had limited effect on the estimation of *R*_t_ for each lineage. We also found that B.1.177 had a similar transmissibility than B.1.1.37 (Student’s t test, two-sided with Holm–Bonferroni adjusted *p* =0.1) (Figure 2D).

**Figure 2.**
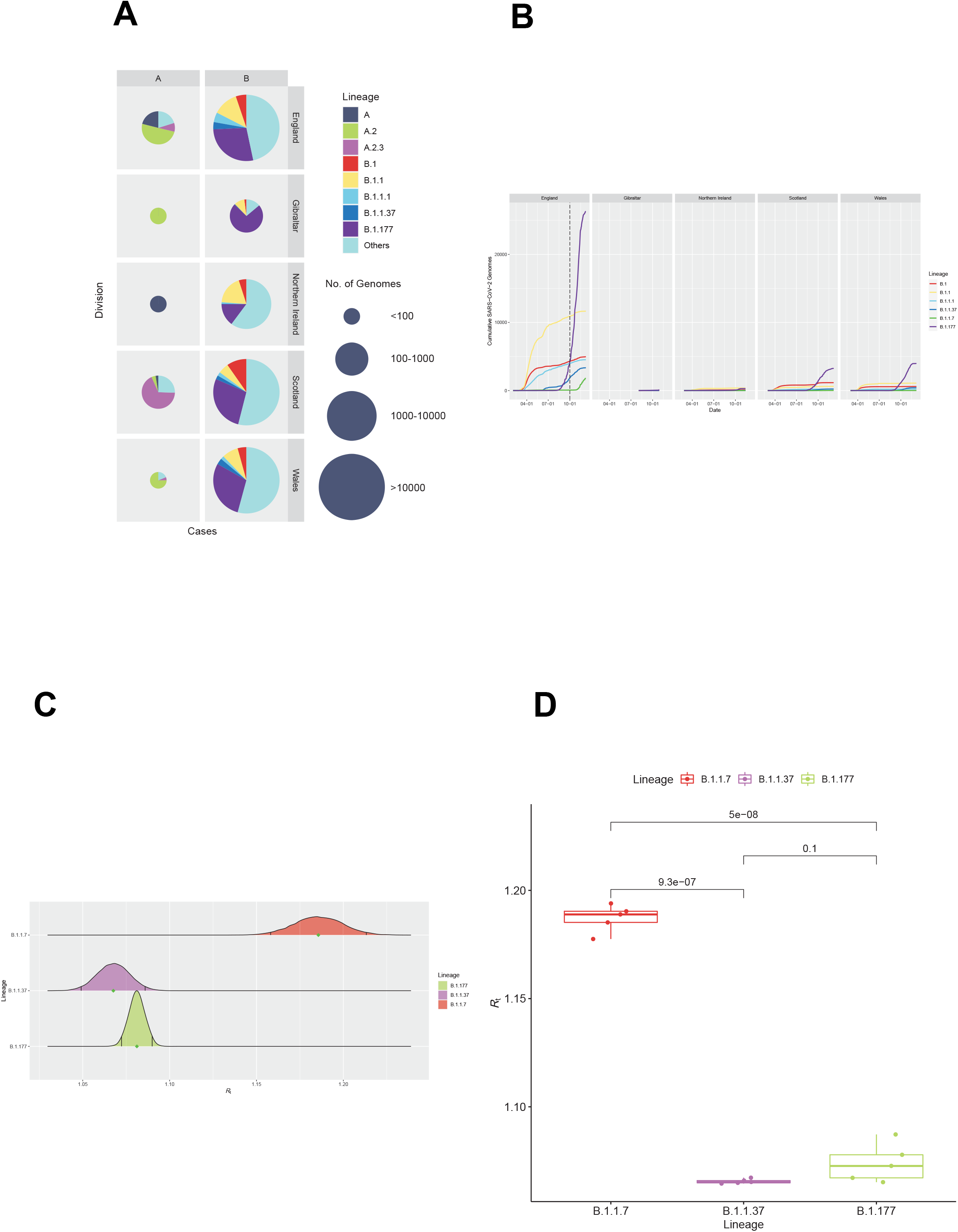
Lineage B of SARS-CoV-2 has a higher transmissibility than lineage A. A. The pie chart of SARS-CoV-2 lineage composition in UK. The circle size was proportion to the number of SARS-CoV-2 genomes. B. The cumulative number of SARS-CoV-2 genomes for each lineage in different region in UK. The dash line indicated the earliest collection date of the data used for estimating the transmissibility for each lineage. C. The distribution of *R*_t_ for each lineage. The black line in each distribution indicated the 95% CI. D. The boxplot of repeated estimation of transmissibility by using 5 independent re-sampling data for each lineage. Upper bound, center, and lower bound of box represent the 75th percentile, the 50th percentile (median), and the 25th percentile, respectively.

### Slightly higher transmissibility for B.1.351 than B.1.1.54 in South Africa

The composition of lineages in South Africa is shown in Figure 3A. Lineage B.1.1.54 was the dominant strain before October 2020. Since then, the dominant strain in South Africa was switched to lineage B.1.351 gradually. According to the accumulation of number of viral genomes from each lineage in South Africa, we could find that only lineage B.1.1.54 and B.1.351 grew exponentially after July 2020 (Figure 3B). In this case, only transmissibility of these two lineages were evaluated during July 2020 to February 2021 in this study, so that the impact of non-pharmaceutical interventions on the estimation of *R*_t_ will be consistent for these two lineages. We could find the *R*_t_ for B.1.351 and B.1.54 during July 2020 and February 2021 were estimated as 1.05 (95% CI 1.044-1.065) and 1.02 (95% CI 1.011-1.034), respectively (Figure 3C). The difference of transmissibility between B.1.351 and B.1.54 was also significant (Student’s t test, two-sided *p*<0.001) (Figure 3D). In this case, isolates from B.1.351 had a slightly higher transmissibility than those from B.1.154.

**Figure 3.**
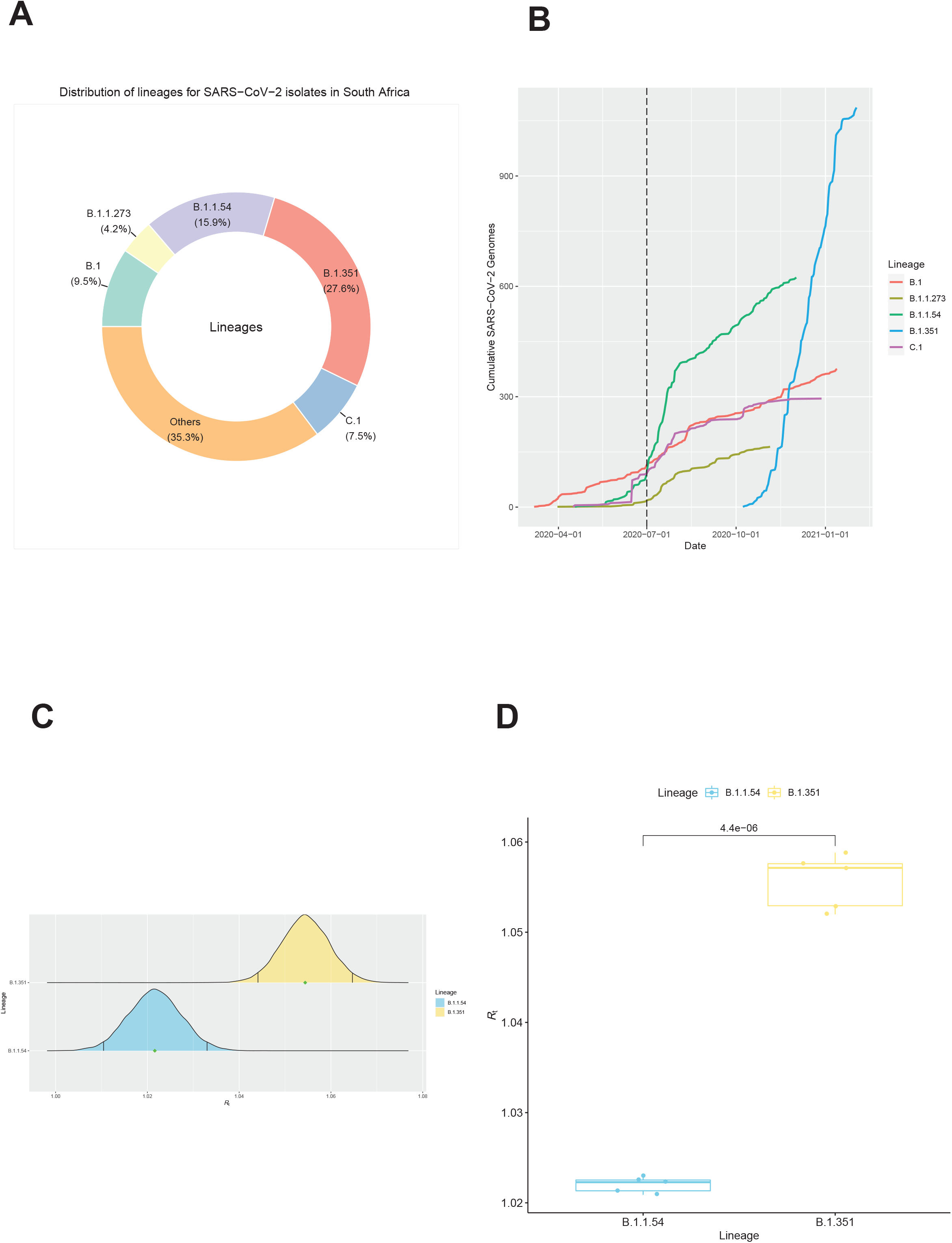
Difference in transmissibility for lineages in South Africa. A. The donut chart of SARS-CoV-2 lineage composition in South Africa. B. The cumulative number of SARS-CoV-2 genomes for each lineage in South Africa. The dash line indicated the earliest collection date of the data used for estimating the transmissibility for each lineage. C. The distribution of *R*_t_ for each lineage. The black line in each distribution indicated the 95% CI. D. The boxplot of repeated estimation of transmissibility by using 5 independent re-sampling data for each lineage. Upper bound, center, and lower bound of box represent the 75th percentile, the 50th percentile (median), and the 25th percentile, respectively.

### P.1 had a slightly higher transmissibility than P.2 in Brazil

The composition of lineages in Brazil is shown in Figure 4A. Lineage B.1.1.33 and B.1.1.28 were the dominated before January 2021. Both of them grew exponentially after their first appearance in Brazil. However, their growth rate has slowed down since July 2020. Since October 2020, two novel lineages (P.1 and P.2) had gradually appeared and had shown exponential growth (Figure 4B). In this case, only transmissibility of these two lineages (P.1 and P.2) were evaluated during December 2020 to February 2021 in this study, so that the impact of non-pharmaceutical interventions on the estimation of *R*_t_ will be consistent for these two lineages. We could find the *R*_t_ for P.1 and P.2 during December 2020 to February 2021 were estimated as 1.07 (95% credible intervals 1.054-1.084) and 1.06 (95% credible intervals 1.049-1.070) (Figure 4C), respectively. The difference of transmissibility between P.1 and P.2 was also significant (Student’s t test, two-sided *p*=0.016) (Figure 4D). In this case, isolates from P.1 had a slightly higher transmissibility than those from P.2.

**Figure 4.**
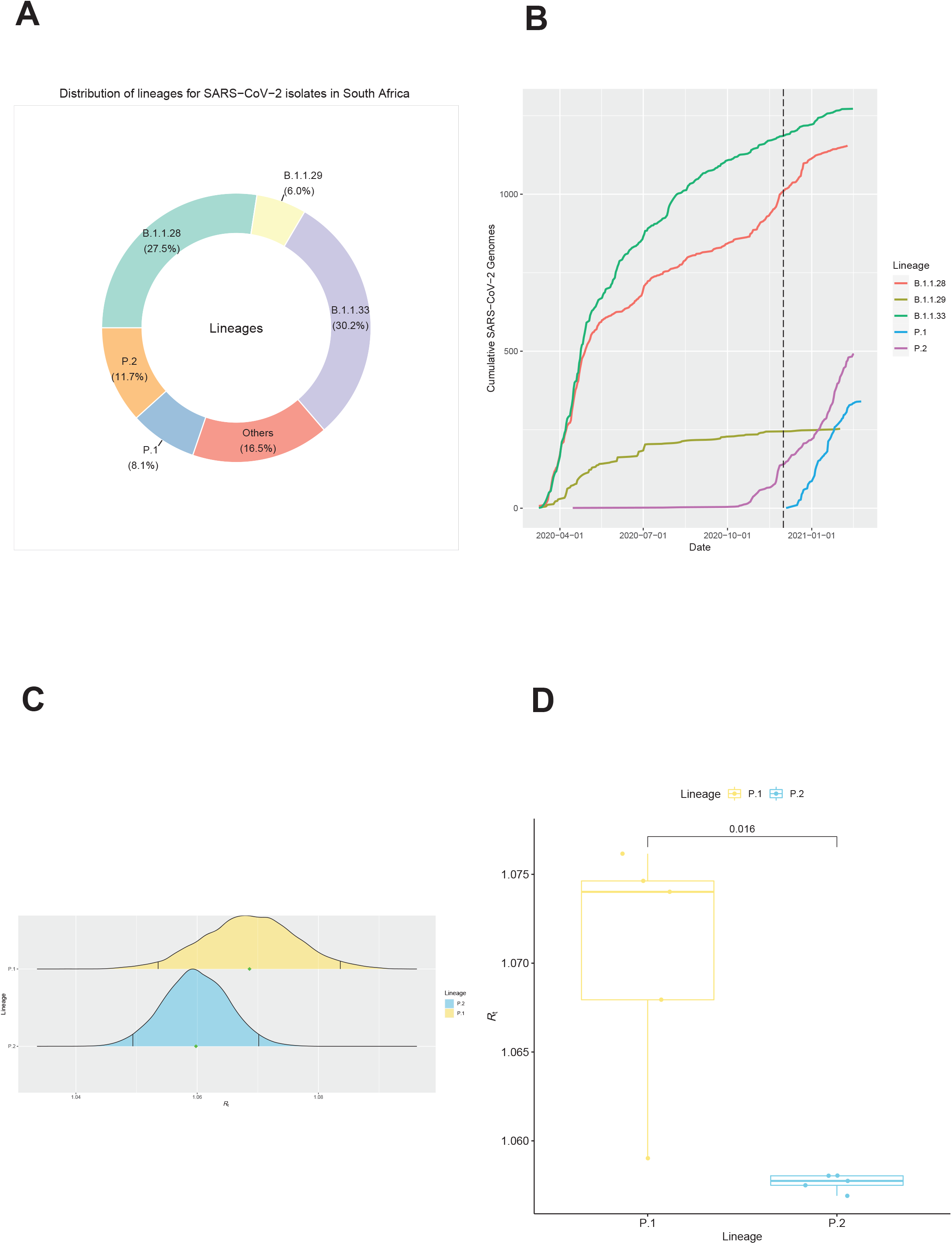
Difference in transmissibility for lineages in Brazil. A. The donut chart of SARS-CoV-2 lineage composition in Brazil. B. The cumulative number of SARS-CoV-2 genomes for each lineage in Brazil. The dash line indicated the earliest collection date of the data used for estimating the transmissibility for each lineage. C. The distribution of *R*_t_ for each lineage. The black line in each distribution indicated the 95% CI. D. The boxplot of repeated estimation of transmissibility by using 5 independent re-sampling data for each lineage. Upper bound, center, and lower bound of box represent the 75th percentile, the 50th percentile (median), and the 25th percentile, respectively.

### B.1.617.2 has a higher transmissibility than other dominant lineages in India

The top five dominant lineages and their corresponding proportion in India are shown in Figure 5A. The B.1.306 was the dominated lineage in India. Since July 2020, several other lineages, like B.1, B.1.36, B.1.36.29, emerged and grew exponentially in India (Figure 5B). B.1.617.1 and B.1.617.2 were detected in India at late 2020, and then they both grew exponentially in India (Figure 5B). Lineage B.1.617.2 has already been considered as VOC by WHO. We also found lineage B.1, B.1.36, B.1.36.29, B.1.617.1, B.1.617.2 grew exponentially after 1^st^ January 2021. In order to reduce the calculation, only data collected after 1^st^ January 2021 were used to perform the further analysis so that the impact of non-pharmaceutical interventions on the estimation of *R*_t_ will be consistent for these lineages. In this case, only these five lineages were used to estimate their *R*_t_. The *R*_t_ was estimated as 1.013 (95% CI 1.006-1.021), 1.018 (95% CI 1.009 1.027), 1.019 (95% CI 1.010-1.027), 1.033 (95% CI 1.026-1.040), 1.123 (95% CI 1.106-1.140) for B.1, B.1.36, B.1.36.29, B.1.617.1, B.1.617.2, respectively (Figure 5C). After 5 independently repeated sampling and followed analysis for each lineage, we found that both B.1.617.1 and B.1.617.2 had significantly higher transmissibility than B.1, B.1.36, and B.1.36.29 (all Student’s t test, two-sided with Holm–Bonferroni adjusted *p*<0.001) (Figure 5D). Furthermore, B.1.617.2 also had a significantly higher transmissibility than B.1.617.1 (Student’s t test, two-sided with Holm–Bonferroni adjusted *p*<0.001). In addition, the transmissibility of both B.1.36, and B.1.36.29 is significantly higher than that of B.1 (both Student’s t test, two-sided with Holm–Bonferroni adjusted *p*<0.001) (Figure 5D). However, similar transmissibility was found between B.1.36 and B.1.36.29 (Student’s t test, two-sided with Holm–Bonferroni adjusted *p*=0.057) (Figure 5D).

**Figure 5.**
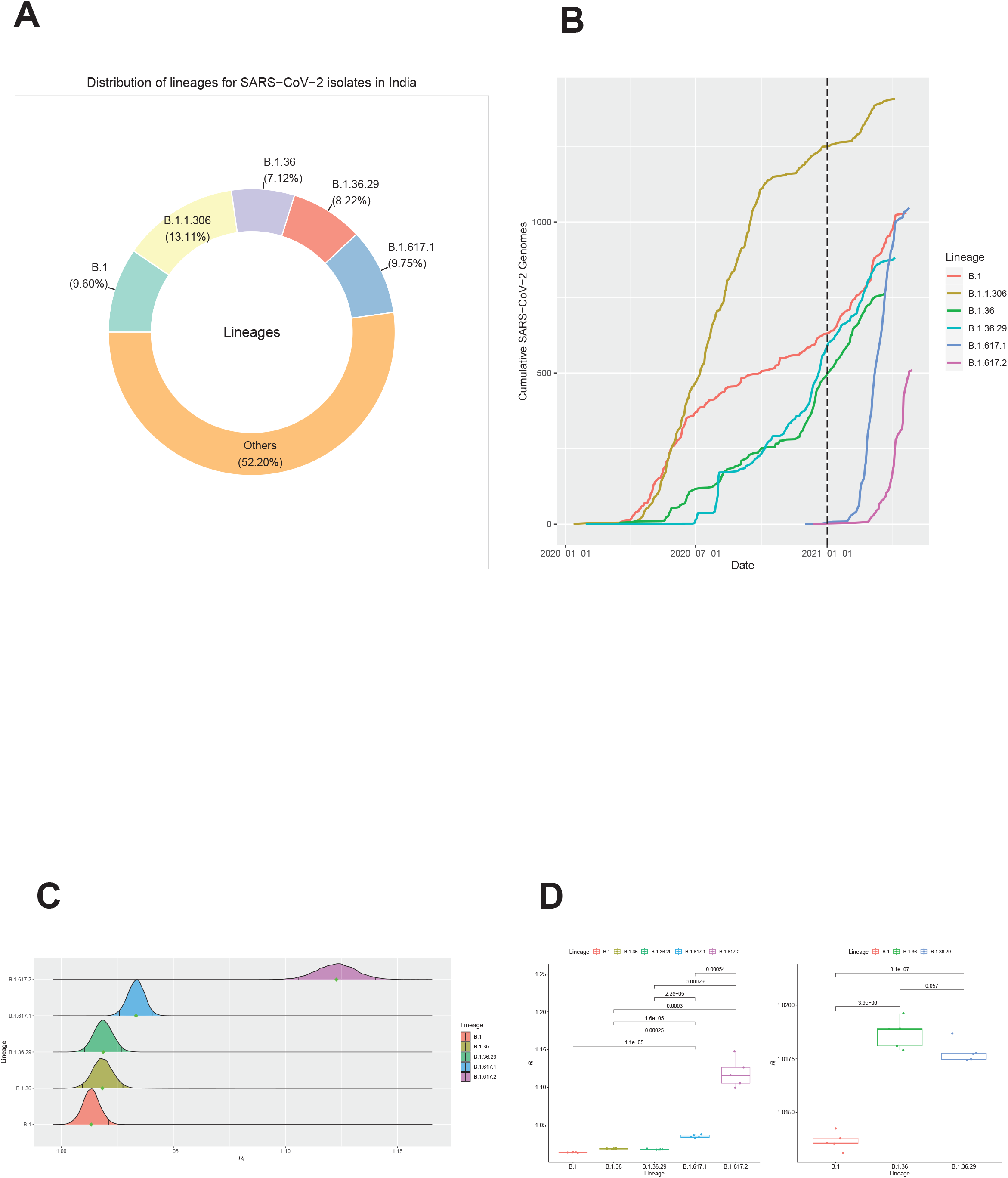
Difference in transmissibility for lineages in India. A. The donut chart of SARS-CoV-2 lineage composition in India. B. The cumulative number of SARS-CoV-2 genomes for each lineage in India. The dash line indicated the earliest collection date of the data used for estimating the transmissibility for each lineage. C. The distribution of *R*_t_ for each lineage. The black line in each distribution indicated the 95% CI. D. The boxplot of repeated estimation of transmissibility by using 5 independent re-sampling data for each lineage. Upper bound, center, and lower bound of box represent the 75th percentile, the 50th percentile (median), and the 25th percentile, respectively.

## Discussion

Assessing the transmissibility of pathogens is essential to tailor prevention and control strategies. As the COVID-19 pandemic spread, several VOC have been found, such as B.1.1.7 (WHO label: Alpha), B.1.351 (WHO label: Beta), P.1 (WHO label: Gamma), and B.1.617.2 (WHO label: Delta) *etc*. The emergence of these VOCs has caused a significant threat to public health. Since vaccination is the key to global containment of the COVID-19 pandemic, a reduced vaccine efficacity against some VOCs would increase the risk of infection in immunized individuals thereby increasing the difficulty of containing the spread of the pandemic. For example, B.1.1.7 has been documented to have reduced neutralization by original strain convalescent and vaccine sera^8-10^. B.1.351 and P.1 also had reduced neutralization by mAbs and sera induced by early SARS-CoV-2 isolates^11-13^ and B.1.351 might also increase the risk of infection in immunized individuals^14^. However, novel VOCs might emerge at any time and anywhere in the future. In order to deal with a novel VOC, it is necessary to quickly evaluate its transmissibility and use this as a basis to determine whether prevention and control strategies need to be adjusted to control the epidemic. A previous study had documented that B.1.1.7 has an advanced transmissibility compared to other lineages circulating in UK (43%-90% with 95% confidence intervals ranging from 38% to 130%)^15^. Another study illustrated that P.1 also had an increased transmissibility by 54%-79% compared to non-P.1 lienage^16^. These results show that different lineage can have different transmissibility. Together with the changes in transmissibility for B.1.351 and B.1.617.2 which had not been previously elucidated, here we estimated the lineage-specific transmissibility for each lineage, especially for these VOCs.

The results show that lineage B has a significantly higher transmissibility than lineage A (Figure 1A). Together with the fact that lineage B was the dominant types of SARS-CoV-2 all over the world, it seems that the high transmissibility of lineage B contributed to the global pandemic to a large extent. However, we also found that the transmissibility for lineage B from Australia and USA differed significantly. Considering the significantly different composition of sub-lineages among these two countries, we speculated that different sub-lineage within lineage B would have different transmissibility. We estimated the transmissibility of VOCs and the dominant lineages with exponential growth during same period in each country, so that the impact of non-pharmaceutical interventions on the estimation of *R*_t_ will be consistent among different lineages. We estimated *R*_t_ for different lineages of SARS-CoV-2 based on genomic data and Bayesian inference under an epidemiological model, and then inferred the mean of offspring distribution (*R*_t_). Since limited variants among each lineage would lead to uncertainty on phylogeny and the estimation of *R*_t_ was solely based on dated-phylogenetic tree, it is necessary to assess how the phylogenetic uncertainty affect the estimation of *R*_t_. The estimation of *R*_t_ from random selected tree from MCMC chain were always lower than for the MCC tree (Figure S1). As the MCC tree is more accurate than to trees sampled in MCMC chains, this result suggested that the uncertainty of the phylogeny would cause an underestimation of the *R*_t_. In this case, the use of MCC tree for estimation of *R*_t_ would reduce the impact of phylogenetic uncertainty on the results as much as possible. In addition, the sampling bias could also affect the phylogeny.

B.1.1.7 had a significant advance in transmissibility than B.1.37 and B.1.177 in UK. The result was consistent with the previous report that B.1.1.7 had a higher transmissibility than other lineages^15^. However, the increase in transmissibility of B.1.1.7 estimated in this study was not as much as previous reported^15^, presumably because the increase of transmissibility of B.1.1.7 was based on comparison to the superimpose state of all other lineages for previous report. On the other hand, the increase of transmissibility of B.1.1.7 was based on comparison to two other lineages (B.1.1.37 and B.1.177) in the UK. Since B.1.1.37 and B.1.177 grew exponentially in the UK, the transmissibility for these two lineages could be higher than those lineages without exponentially growth. In this case, the increase of B.1.1.7 was not as much as higher than previous report^15^. The transmissibility of B.1.1.7 has indeed increased and is in line with the results from other reports, which further proves the accuracy of our method. We also found that P.1 had a higher transmissibility than P.2 in Brazil, and B.1.351 had a higher transmissibility than B.1.1.54 in South Africa. However, the extent of increased transmissibility for P.1 and B.1.351 against to other dominant lineages with exponential growth was not as much as for B.1.1.7. In India, we found that both B.1.617.1 and B.1.617.2 had significant increase in transmissibility compared to other lineages with exponential growth. Furthermore, B.1.617.2 also had a significantly higher transmissibility than B.1.617.1. In addition, B.1.36 and B.1.36.29 had similar transmissibility, both higher than B.1.

These results indicated that different lineages of SARS-CoV-2 have different transmissibility, with some differences being more significant than others. The transmissibility of four types of VOCs also increased to varying degrees. All countries should formulate corresponding prevention and control policies for these VOCs to avoid large-scale outbreaks in their countries.

## Methods

### Data collection, selection and pre-processing

The transmission could be significantly affected by the stringent prevention and control strategies. Only data collected before the implementation of stringent epidemic control measures in each country were used for lineages A and B, so as to minimize the impact of prevention and control strategies on the estimation of *R*_t_, and reflect the real situation at the same time. SARS-COV-2 genomic sequences were download from GISAID several times (data for estimating lineage A and B was downloaded at 9^th^ April 2020, data for UK was downloaded at 21^st^ December 2020, data for South Africa and Brazil was downloaded at 16^th^ March 2021, data for India was downloaded at 13^th^ May 2021). Before estimating transmissibility of lineage A and B during the early phase of COVID-19 pandemic, we first filtered data. Only those viral genomes collected before the implementation of national non-pharmaceutical interventions would be included in the analysis. In addition, those countries that include lineage A and B, and the number of completely viral genomes within each lineage ≥80 would be included in the subsequent analysis. Since only the United States and Australia met the above criteria, the estimation of the transmissibility of lineage A and B was only based on the data of these two countries. The cut-off dates for the collection time in the USA and Australia are 20^th^ and 25^th^ January 2020, respectively, as there were no nationwide epidemic prevention measures were implemented before the date. Due to the high volume of genomic data from sub-lineages in UK, South Africa, Brazil, and India, the amount of calculation would be too large, especially for reconstruction of dated phylogeny. In this case, we filtered and also sub-sampled the data for datasets from each sub-lineage. First, the viral genomes of patients who had not had a history of international travel are retained, according to their epidemiological data. Second, the viral genomes should also meet the criteria as follow: length ≥29 KB, and the ratio of N in the genome ≤1%. Third, based on the collection date, if more than 10 genomes were available in a specific date, we randomly select 10 of them, otherwise all genomes would be included. Genomic sequences were aligned using Mafft v7.310^17^. Then, we trimmed the uncertain regions in 3′ and 5′ terminals and also masked 30 sites (Supplementary Table 1) that are highly homoplastic and have no phylogenetic signal as previous noted (https://virological.org/t/issues-with-sars-cov-2-sequencing-data/473).

### Reconstruction of dated phylogeny

As recombination could impact the evolutionary signal, we searched for recombination events in these SARS-CoV-2 genomes using RDP4^18^. No evidence for recombination was found in our dataset. We used jModelTest v2.1.6^19^ to find the best substitution model for each dataset from different countries according to the Bayesian Information Criterion. The best substitution model for each dataset were listed in Supplementary Table 2. The list of genomic sequences used in this study were provided in Supplementary Table 3. The list of genomic sequences used in this study were openly shared via the GISAID initiative^20^. We then used the Bayesian Markov Chain Monte Carlo (MCMC) approach implemented in BEAST v1.10.4^21^ to derive a dated phylogeny for SARS-CoV-2. Three replicate runs for each 100 million MCMC steps, sampling parameters and trees every 10,000 steps. For data from lineage A and B in USA and Australia during the early phase of COVID-19, the estimation of the most appropriate combination of molecular clock and coalescent models for Bayesian phylogenetic analysis was determined using both path-sampling and stepping-stone models^22^. The model comparison result for datasets from lineage A and B in USA and Australia were listed in Supplementary Table 4. In order to reduce the amount of calculation, we assumed that data from sub-lineages followed a strict molecular clock and with an exponential population growth tree prior, as genomic sequences used in each dataset were all from the same sub-lineage and they all had an exponential growth. Tracer 1.7.1^23^ was then used to check the convergence of MCMC chain (effective sample size >200) and to compute marginal posterior distributions of parameters, after discarding 10% of the MCMC chain as burn-in. Bayesian evaluation of temporal signal (BETS)^24^ was used to evaluate the temporal signal in each dataset. BETS relies on the comparison of marginal likelihoods of two models: the heterochronous (with tip date) and isochronous (without tip date) models. Analyses were performed with at least three independent replicates of 100 million MCMC steps each, sampling parameters and trees every 10,000 steps with the best substitution model and most appropriate combination of molecular clock and coalescent models determined above for each dataset. The marginal likelihoods were estimated by PS. The Bayes factor (BF) was then calculated based on the likelihoods of two models (heterochronous and isochronous). If the log BF >30 (heterochronous model against isochronous model), it indicated there was sufficient temporal signal in this dataset. For dataset without sufficient temporal signal, we specified a clock rate following uniform distribution ranging from 0.0004 to 0.0012 with a mean of 0.0008, otherwise we specified a noninformative continuous-time Markov chain (CTMC) reference prior. The log BF for each dataset was listed in Supplementary Table 5.

### Estimation of transmissibility using partially sampled viral genomic sequences

As viral genomes were incompletely sampled and the pandemic is currently ongoing, TransPhylo v1.4.4^25^ was used to infer the transmission tree using the dated phylogeny generated above as input. The generation time (i.e., the time gap from infection to onward transmission, denoted as G) of COVID-19 was previously estimated as 7.5 ± 3.4 days^26^ and we used these values to compute the shape and scale parameter of a gamma distribution of G using the R package epitrix^27^. This parameter was used when estimating the transmissibility of lineages A and B in USA and Australia during the early phase of COVID-19 pandemic. However, it was reported that the G was shorten over time by nonpharmaceutical interventions^28^. In this case, we used 4.8±1.7 days^29^ estimated by previous study as G when estimating the transmissibility of sub-lineages in UK, South Africa and Brazil. The distribution of sampling time (*i*.*e*. the time gap from infection to detection and sampling) was set equal to the distribution of generation time. We performed the TransPhylo analysis with 100,000 iterations estimating the the offspring distribution (which represents the number of secondary cases caused by each infection). The *R*_t_ then could be inferred as the median of the offspring distribution. All results were generated after discarding the first part of the MCMC chains as burn-in. The MCMC mixing and convergence was assessed based on the effective sample size of each parameter (>200) and by visual examination of the MCMC traces. The effective sample size and value of *R*_t_ for each dataset was listed in Supplementary Table 6.

### Evaluating the robustness of the estimation

Since dated phylogeny was used to estimate the transmissibility for each lineage, we should test whether and how the phylogenetic uncertainty and sampling bias affect the estimation. We first tested how the phylogenetic uncertainty affect the result, because only MCC tree was used to estimate the transmissibility. We used data from our previous study^30^. Ten dated phylogenetic trees were randomly selected from the MCMC chains. The parameter setting was the same as previous study description. In addition, the sampling bias was also a key factor affecting the phylogenetic uncertainty. In order to test the robustness of the estimation of *R*_t_, we also repeatedly randomly sub-sampled the data five times for each dataset and then performed the same analysis.

## Data Availability

All data used in this manuscript is available in GISAID

## Supplementary Information

Figure S1. The 95% CI distribution of *R*_t_ using MCC tree and ten randomly selected trees from the MCMC chains.

Supplementary Table 1. List of 30 masked sites in SARS-CoV-2genome.

Supplementary Table 2. The best substitution model for dataset from each country.

Supplementary Table 3. The acknowledgement table of viral genomes used in this study.

Supplementary Table 4. Log-marginal likelihood estimates from model selection by using the path-sampling (PS) and stepping-stone (SS) approaches for lineage A and B.

Supplementary Table 5. Bayesian evaluation for the temporal signal of dataset from each dataset.

Supplementary Table 6. The estimation of *R*_t_ and corresponding effective size of each dataset.

